# Dynamic reconfiguration of aperiodic brain activity supports cognitive functioning in epilepsy: a neural fingerprint identification

**DOI:** 10.1101/2024.04.19.24305881

**Authors:** Emahnuel Troisi Lopez, Marie-Constance Corsi, Alberto Danieli, Lisa Antoniazzi, Marianna Angiolelli, Paolo Bonanni, Pierpaolo Sorrentino, Gian Marco Duma

**Author notes:** Corresponding author: Gian Marco Duma. Authors contributed equally as first authors (Emahnuel Troisi Lopez, Marie-Constance Corsi). Authors contributed equally as last authors (Pierpaolo Sorrentino, Gian Marco Duma).

## Abstract

**Background:** Temporal lobe epilepsy (TLE) is characterized by alterations of brain dynamic at large scale associated with altered cognitive functioning. Interindividual variability of brain activity is a source of heterogeneity in this disorder. Here, we aimed at analyzing dynamical reconfiguration of brain activity, using the neural fingerprint approach, to delineate subject-specific characteristics and their cognitive correlates in TLE.

**Methods:** We collected 10 minutes of resting state electroencephalography (128 channels), free from epileptiform activity, from 68 TLE patients and 34 healthy controls. The functional network was defined by the spatio-temporal spreading, across cortical regions, of aperiodic bursts of activations (neuronal avalanches). This metric allowed encapsulating brain reconfiguration patterns into the avalanche transition matrix (ATM). We used a neural fingerprint approach to differentiate across controls and patients diagnosis, linking altered brain dynamic with cognitive outcome.

**Findings:** Patients’ brain dynamics were more stereotyped as compared to controls. The neural fingerprint using ATMs differentiated, in a data-driven fashion, patients with respect to healthy controls, being sensitive not only to the pathology but also to the subtype (unilateral vs. bilateral TLE). Notably, in unilateral TLE patients, a better memory performance was associated with a larger similarity of the brain dynamic configuration with controls.

**Interpretation:** TLE is characterized by reduced variability and more stereotyped brain dynamics, implicating widespread alterations across the brain. These alterations correlate with cognitive function in patients with unilateral TLE. This study underscores the utility of brain fingerprinting in elucidating disease-specific brain dynamics, offering novel metrics for personalized patient care.

## 1. Introduction

In the last decades, understanding epilepsy as a network disease has proved successful in the conceptualization of its pathophysiology (Bartolomei et al., 2017; Corona et al., 2023). In particular, alterations of brain dynamics on the large-scale have been identified in several epilepsy types, and they are related both to clinical and neuropsychological outcomes (Courtiol et al., 2020; Duma et al., 2021). Amongst the epilepsy types, temporal lobe epilepsy (TLE) is the most frequent drug-resistant focal epilepsy. A proportion of patients with TLE displays bilateral (simultaneous and/or independent) temporal ictal involvement, a condition defined as bilateral temporal lobe epilepsy (BTLE) (Aghakhani et al., 2014). Some studies have described distinctive clinical-anatomo-electrophysiological features of BTLE as compared to unilateral TLE (UTLE), suggesting that BTLE may be considered a relatively specific condition within the TLE spectrum (Di Vito et al., 2016; Didato et al., 2015). From a neurocognitive perspective, TLE has been associated with impairment in different cognitive domains, including memory, language, attention, and executive functions (Ives-Deliperi & Butler, 2021; Jokeit et al., 2000). Converging evidence has shown a link between the disruption of large-scale functional organization in TLE and cognitive performance (Duma et al., 2022; Girardi-Schappo et al., 2021; He et al., 2018). More specifically, recent findings suggested that patients with BTLE are characterized by worse neuropsychological performance as compared to UTLE (Baggio et al., 2023). Cognitive functions rely on the coordinated interactions amongst multiple brain areas over time. In fact, in the healthy brain, the capability of reorganization of large-scale functional properties (Zalesky et al., 2014) has been related to cognitive proficiency (Braun et al., 2015; Kao et al., 2020; Mattar et al., 2015). The dynamics of the reconfigurations over time contain enough information to unambiguously identify individuals, representing a subject-specific neural fingerprint (da Silva Castanheira et al., 2021). Neurological diseases can induce a loss of the neural fingerprint, which has been deployed as a clinical biomarker (Cipriano et al., 2023; Sorrentino, Rucco, et al., 2021; Troisi Lopez et al., 2023). Concerning epilepsy, a dysregulation of the whole-brain dynamics, captured by scalp-electroencephalography (EEG) derived microstates, has been identified as a potential neural signature differentiating not only patients from controls, but also UTLE from BTLE (Baldini et al., 2024). Indeed, microstates are stable configurations of topographical EEG maps related to an underlying functional organization on the large scale (Michel & Koenig, 2018). In this perspective, the present study aims at exploiting dynamical features captured by the EEG signals in order to define the neural fingerprint of patients with epilepsy as compared to controls. Additionally, the second aim of this work is to identify potential biomarkers to provide evidence about whether BTLE constitutes a distinct nosological entity with respect to TLE. To this purpose, we focused on the topography of the spreading of aperiodic perturbations across the whole-brain (Rucco et al., 2020; Sorrentino, Petkoski, et al., 2022), namely the neuronal avalanches (NA). The NA represent the aperiodic bursts of brain activities spreading over the large scale (Romano et al., 2023; Sorrentino, Seguin, et al., 2021). Converging evidence has shown that functional connectivity is driven by these aperiodic bursty components (Sorrentino, Rabuffo, et al., 2022; Zamani Esfahlani et al., 2020). Specifically, here we exploited avalanche transition matrices (ATMs) to track the propagation of the aperiodic bursts across brain regions (Sorrentino, Seguin, et al., 2021). The ATMs are a TLE-sensitive measure, which provides information about functionally altered regions, as well as the relationship of the disrupted brain dynamics and the morphological configuration of the gray matter in patients with epilepsy (Duma et al., 2023, 2024). Moreover, the ATMs are optimally suited to capture fingerprinting, as compared to classical functional connectivity measures (Corsi et al., 2023; Sorrentino et al., 2023). Here, we exploited ATMs to capture the neural fingerprint of patients with TLE vs. a control group. To this purpose, we recorded 10 minutes of resting state activity with high-density EEG (hdEEG, 128 channels) from which we performed electrical source imaging. No seizures were recorded during the resting state. However, we purposely excluded interictal epileptic discharges (IEDs) to investigate if the basal configuration of the brain, irrespective of epileptiform activities, could provide enough information to differentiate between individuals with epileptic conditions (UTLE vs. BTLE), and healthy controls. We hypothesize that the patients may express increased differentiability as compared to controls, given the alteration observed in their dynamical activity in previous studies (Duma et al., 2021, 2022, 2023). In other words, the presence of the alterations in TLE provokes changes in the dynamics, such that “healthy”, optimally flexible dynamics are lost, in a way that is specific to each patient. As a consequence, we expect the patients to be less similar to each other, as compared to how much the healthy subjects are similar to each other. Conversely, the impoverished, less flexible dynamics of each patient would lead to activities that are more similar over time, within each patient. We expect this pattern to be more pronounced in BTLE patients as compared to UTLE patients, to be able to differentiate between these two populations (Baldini et al., 2024; Didato et al., 2015). Finally, we hypothesized that alterations in brain fingerprinting might capture suboptimal cognitive functioning. In particular, considering that dynamic functional flexibility is one of the scaffolding elements of cognitive function, we expect to detect larger similarity of the reconfiguration patterns of activity, as compared to healthy controls, in patients with a better cognitive outcome.

## 2. Methods

### 2.1. Participant

We retrospectively enrolled 72 patients with temporal lobe epilepsy, who underwent high-density electroencephalography (hdEEG) for clinical evaluation between 2018-2022 at the Epilepsy and Clinical Neurophysiology Unit, IRCCS Eugenio Medea in Conegliano (Italy). The diagnostic workflow included clinical history and examination, neuropsychological assessment, long-term surface Video EEG (32 channels) monitoring, high-density EEG (hdEEG) resting-state recording, magnetic resonance imaging (MRI) of the brain, and positron emission tomography (PET) as an adjunctive investigation in selected cases. The diagnosis of temporal lobe epilepsy was established according to the ILAE guidelines. A number of 4 patients received invasive surgery before the hdEEG recording. For this reason, we reduced the final sample to 68 (31 left-TLE; 17 right-TLE; 20 bilateral TLE) (whole sample mean age = 41.40 [SD = 17.11]; 33 females). A description of patients’ demographic and clinical characteristics is provided in Table 1. The control group was composed of 35 healthy participants with no history of neurological or psychiatric disorders (mean age = 34.92 [SD = 9.22]; 25 females). Patients signed the informed consent for the study. The study protocol was conducted according to the Declaration of Helsinki and approved by the Comitato Etico Area Nord Veneto (number protocol: 0001878/24). A description of patients’ demographic and clinical characteristics is provided in Table 1.

**Table 1.**
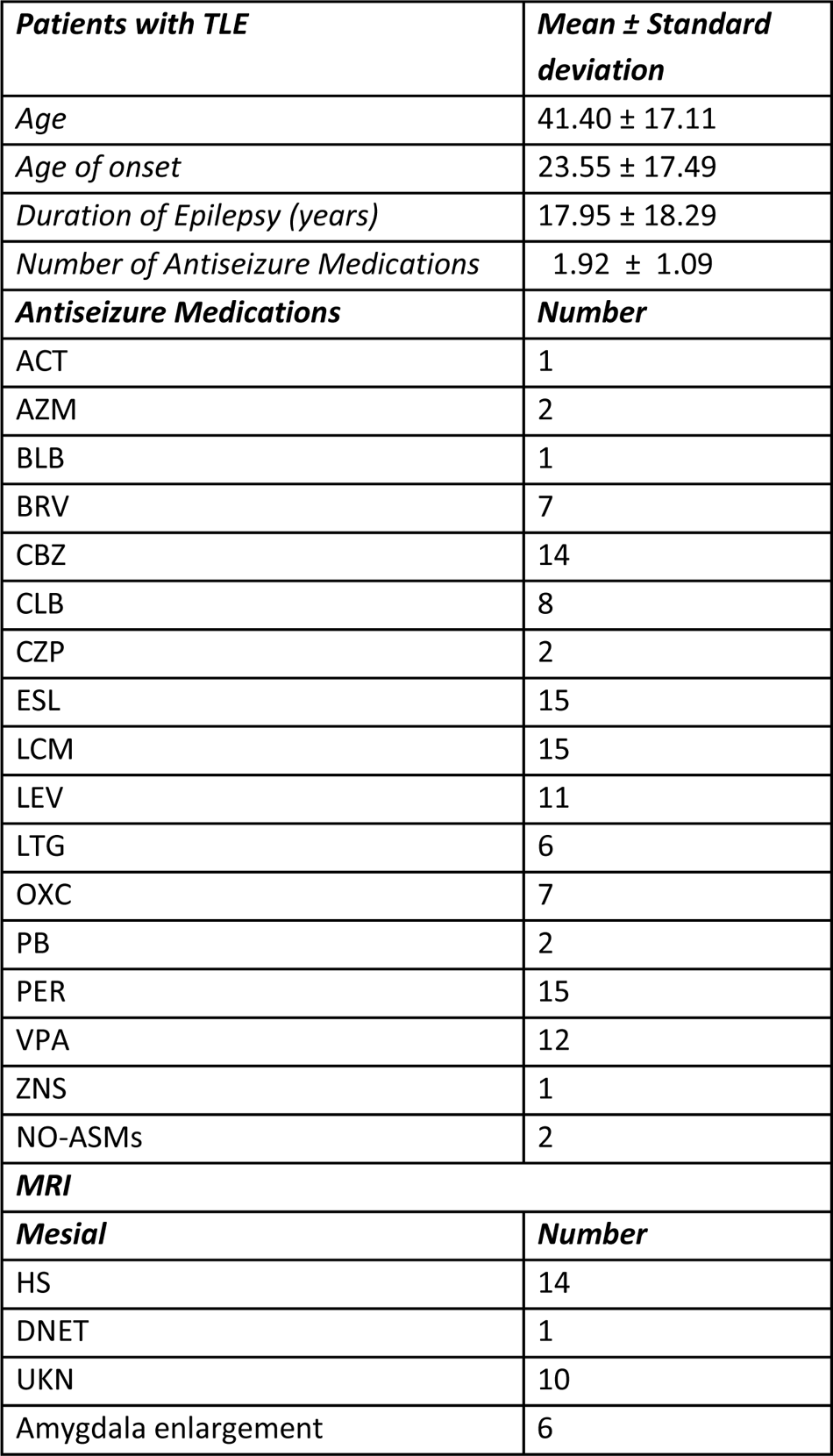
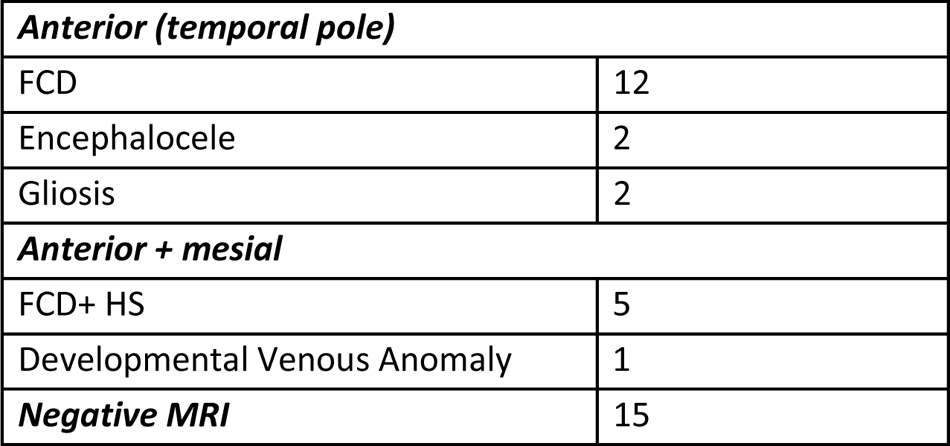
Demographic and clinical characteristics of the patients with temporal lobe epilepsy. The table describes the demographic and clinical characteristics of the patients with temporal lobe epilepsy. MRI abnormalities are reported by sublobar localization. The continuous variables are reported as mean ± SD. Antiseizure medication abbreviations: ACT, acetazolamide; AZM, acetazolamide; BRV, brivaracetam; CBZ, carbamazepine; CLB, clobazam; CZP, clonazepam; ESL, eslicarbazepine; LCM, lacosamide; LEV, levetiracetam; LTG, lamotrigine; OXC, oxcarbazepine; PB, phenobarbital; PER, perampanel; VPA, valproic acid; ZNS, zonisamide; NO-ASMs, no pharmacological treatment. Abbreviation of the identified anomalies on the MRI: FCD, focal cortical dysplasia; HS, hippocampal sclerosis; DNET, dysembryoplastic neuroepithelial tumours; UKN, unknown.

| ***Patients with TLE*** | ***Mean ± Standard deviation*** |
| --- | --- |
| *Age* | 41.40 ± 17.11 |
| *Age of onset* | 23.55 ± 17.49 |
| *Duration of Epilepsy (years)* | 17.95 ± 18.29 |
| *Number of Antiseizure Medications* | 1.92 ± 1.09 |
| ***Antiseizure Medications*** | ***Number*** |
| ACT | 1 |
| AZM | 2 |
| BLB | 1 |
| BRV | 7 |
| CBZ | 14 |
| CLB | 8 |
| CZP | 2 |
| ESL | 15 |
| LCM | 15 |
| LEV | 11 |
| LTG | 6 |
| OXC | 7 |
| PB | 2 |
| PER | 15 |
| VPA | 12 |
| ZNS | 1 |
| NO-ASMs | 2 |
| ***MRI*** |  |
| ***Mesial*** | ***Number*** |
| HS | 14 |
| DNET | 1 |
| UKN | 10 |
| Amygdala enlargement | 6 |
| ***Anterior (temporal pole)*** |  |
| FCD | 12 |
| Encephalocoele | 2 |
| Gliosis | 2 |
| ***Anterior + mesial*** |  |
| FCD+ HS | 5 |
| Developmental Venous Anomaly | 1 |
| ***Negative MRI*** | 15 |

### 2.2. Resting State EEG recording

The hdEEG recordings were obtained using a 128-channel Micromed system referenced to the vertex. Data was sampled at 1,024 Hz and the impedance was kept below 5kΩ for each sensor. For each participant, we recorded 10 minutes of closed-eyes resting state while comfortably sitting on a chair in a silent room.

### 2.3. EEG pre-processing

Signal preprocessing was performed offline via EEGLAB 14.1.2b 22 (Delorme & Makeig, 2004). The continuous EEG signal was downsampled at 250 Hz and then bandpass-filtered (0.1 to 45 Hz) with a Hamming windowed sinc finite impulse response filter (filter order = 8250). The visual inspection was performed to identify interictal epileptiform discharges (IEDs) by the clinicians. The signal was then segmented into 1-sec-long epochs and epochs containing IEDs were removed. We purposely removed epochs containing IEDs since we wanted to focus on the intrinsic brain functional organization, independent of epileptiform activities. The epoched data underwent automated bad-channel and artifact detection algorithm using the TBT plugin implemented in EEGLAB. This algorithm identified the channels that exceeded a differential average amplitude of 250μV and marked those channels for rejection. Channels that were marked as bad in more than 30% of all the epochs were excluded. Additionally, epochs having more than 10 bad channels were excluded. We automatically detected possible flat channels with the Trimoutlier EEGLAB plug-in within the lower bound of 1μV. We rejected an average of 55.04 ± 52.96 (SD) epochs related to IED and artifacts. This preprocessing analysis pipeline has been applied by our group in previous studies investigating neuronal avalanches in epilepsy with resting state EEG activity (Duma et al., 2023, 2024). Data cleaning was performed with independent component analysis, using the Infomax algorithm as implemented in EEGLAB. The resulting 40 independent components were visually inspected and those related to eye blinks, eye movements, muscle, and cardiac artifacts were discarded. An average of 9.89± 4.75 (SD) components were removed. The remaining components were then projected back to the electrode space. Finally, bad channels were reconstructed with the spherical spline interpolation method (Perrin et al., 1989). The data were then re-referenced to the average of all electrodes. At the end of the data preprocessing, each subject had at least 6 minutes of artifact-free signal.

### 2.4. Cortical Source modeling

We used the individual anatomy MRI to generate individualized head models for the patients with TLE. The anatomic MRI for source imaging consisted of a T1 isotropic three-dimensional (3D) acquisition. For twelve patients and for the control group we used the MNI-ICBM152 default anatomy (Evans et al., 2012) from Brainstorm (Tadel et al., 2011), since the 3D T1 MRI sequences were not available. The MRI was segmented into skin, skull, and gray matter using the Computational Anatomy Toolbox (CAT12) (Dahnke et al., 2013). The resulting individual surfaces were then imported in Brainstorm, where three individual surfaces adapted for Boundary Element Models (BEM) were reconstructed (inner skull, outer skull and head) and the cortical mesh was downsampled at 15,002 vertices. The co-registration of the EEG electrodes was performed using Brainstorm by projecting the EEG sensor positions on the head surface with respect to the fiducial points of the individual or the template MRI (Fig. 1A). We applied manual correction of the EEG cap on the individual anatomy whenever needed, prior to projecting the electrodes on the individual head surface. We then derived an EEG forward model using the 3-shell BEM model (conductivity: 0.33, 0.165, 0.33 S/m; ratio: 1/20) estimated using OpenMEEG method implemented in Brainstorm (Gramfort et al., 2010; Kybic et al., 2005). Finally, we used the weighted minimum norm (Hämäläinen & Ilmoniemi, 1994) imaging as the inverse model, with the Brainstorm’s default parameters setting.

**Figure 1.**
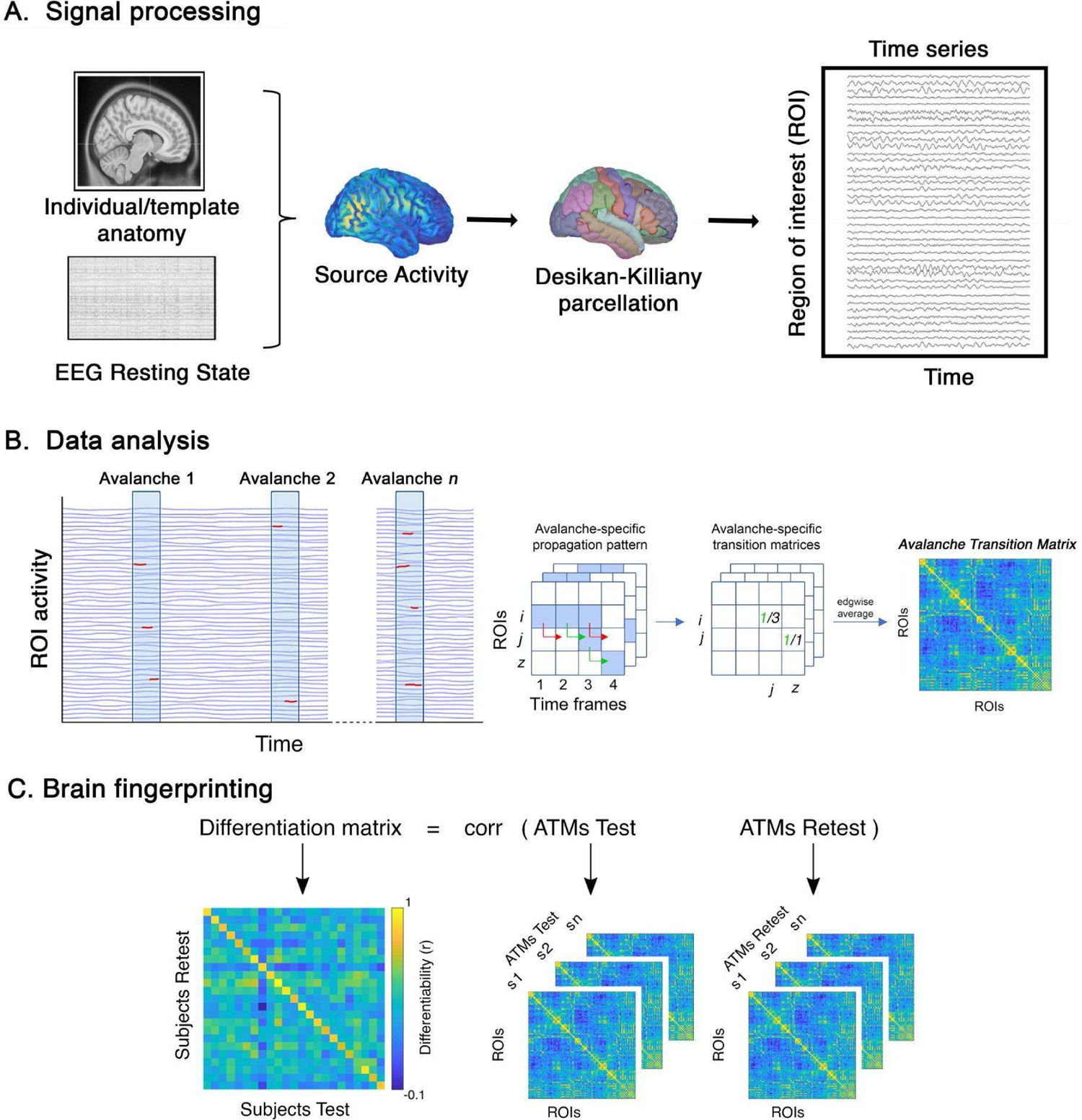
Processing and analysis pipelines. **A** displays the signal processing pipeline from resting state EEG to the source reconstruction and downsampling to a set of 68 regions of interest (ROI) using the Desikan-Killiany cortical parcellation to extract ROIs time series. **B** displays the data analysis pipeline: neuronal avalanches individuation; activity propagation; avalanche transition matrix (ATM). **C** displays the fingerprinting analysis: test and re-test ATMs of each group were correlated, separately, obtaining a differentiation matrix for each group.

### 2.5. Brain dynamics

To explore the dynamics of brain activity, we derived “neuronal avalanches” from the time series reconstructed at its source (Fig. 1B). Firstly, for a fair comparison we used the same duration for each recording of each participant. For a fingerprint analysis we needed two recordings (test and retest) for each individual, hence each recording was composed of a time series of 100 seconds. Then, we discretized the time series for each region of interest by calculating the z-score as follows:

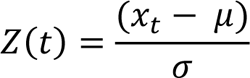

where x is the signal, μ is the average value of the signal across time, and σ is its standard deviation.

Subsequently, we detected both positive and negative excursions surpassing a specified threshold, as:

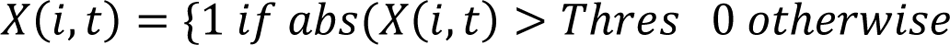

Specifically, the analyses were performed setting the threshold at 2.8 standard deviations, and then repeated at 2.6 and 3 standard deviations, to check that the results were not dependent on a single specific threshold. A neuronal avalanche begins when, in a sequence of contiguous time bins, at least one ROI is active (i.e., above threshold), and ends when all ROIs are inactive (Sorrentino, Seguin, et al., 2021). Then, to ensure that we were observing a system operating in a near-critical regime, we calculated the branching ratio (i.e., a measure that characterizes the division or divergence of pathways within a structure or process), that in systems operating at criticality typically displays value ∼1. Specifically, the branching ratio was determined by geometrically averaging the ratio of the number of events (activations) between subsequent time bins and the current time bin, over all time bins, and then averaging it across all avalanches, as:

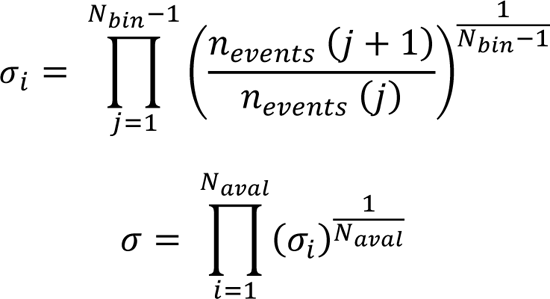

Where σi is the branching parameter of the i-th avalanche in the subject, Nbin is the total amount of bins in the i-th avalanche, Naval is the total number of avalanches in the dataset. Then, for each avalanche n, the transition matrix AvalATM (n) was defined as:

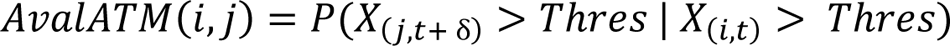

where the element (i, j) represents the probability that region j is active at time t+ẟ, given that region i was active at time t, where ẟ∼3ms. The ATMs were averaged within each participant, as:

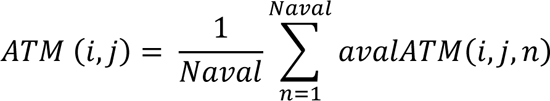

and finally symmetrized. Introducing a time delay diminishes the likelihood that our findings can be easily attributed to field spread. Field spread refers to the simultaneous detection of multiple sources by various sensors, leading to spurious zero-lags correlations in the recorded signals. For a detailed analysis of field spread in this dataset, please consult (Sorrentino et al., 2021c).

### 2.6. Fingerprint analysis

We based our fingerprinting analysis on the brain dynamics by the means of ATMs, similarly to (Sorrentino et al., 2023). Initially, our goal was to construct an Identifiability Matrix (IM) following the methodology outlined by Amico and Goñi (Amico & Goñi, 2018) (Fig. 1C). The IM organizes participants into rows and columns, with entries representing Pearson’s correlation coefficients between the test and retest ATMs for each participant. The IM encapsulates information on self-similarity (Iself, found on the main diagonal elements), indicating the comparison of test and retest ATMs for the same participant. Additionally, it includes the similarity of each subject with others (Iothers, off-diagonal elements), signifying the resemblance between different individuals of the same group. Then, by computing the difference between the Iself and Iothers, we derive the differential Identifiability (Idiff) (Amico & Goñi, 2018; Sorrentino, Rucco, et al., 2021) that provides an estimate of the fingerprint level within a specific group. Lastly, by correlating the test-retest ATMs of healthy individuals and patients, we can obtain the Iclinical score (referred to as “clinical identifiability” or “clinical fingerprint”). This score reflects the similarity of a patient in comparison to healthy subjects.

For a more in-depth understanding, please consult (Sorrentino, Rucco, et al., 2021).

### 2.7. Edges’ stability

Then, we wanted to assess the stability of pathways of activation represented by the edges of the ATMs. Building upon earlier work on identifiability (Amico & Goñi, 2018), we utilized the intraclass correlation coefficient (ICC) (Koch, 2004) to assess the edge-wise reliability of individual connectomes. Edges exhibit high ICC values when they consistently demonstrate similar levels of synchronization across test-retest sessions.

### 2.8. Neuropsychological assessment

All the patients (UTLE and BTLE) underwent a neuropsychological assessment focusing on memory, attention/executive functions and intelligence. Short-term memory (STM) was investigated with Digit Span Test and Corsi block tapping test (Wechsler, 1945) while long term memory (LTM) was studied via Rey–Osterrieth Complex Figure Test (ROCFT) (Caffarra et al., 2002) and Rey Auditory Verbal Learning Test (Carlesimo et al., 1996). Attention and executive functions were evaluated with the Trail Making Test (TMT) (Giovagnoli et al., 1996). Specifically, we included in the analysis both the part A and B as a measure of motor speed and shifting capabilities, respectively. Finally, we used the total IQ of the WAIS-IV (Wechsler, 1981) or WISC-IV (Orsini et al., 2015) scales as a global intelligence measure. Table 2 shows the descriptive statistics of the neuropsychological scores.

**Table 2.** Neuropsychological scores. The present table shows the mean value across groups of the neuropsychological performance. Abbreviation: IQ = intelligent quotient; RAVLT = Rey auditory verbal learning test; ROCF = Rey–Osterrieth complex figure test; TMT A/B = trail making test A/B

| Test | Score (mean ± standard dev) |
| --- | --- |
| *Digit Span* | 5.63 ± 1.12 |
| *Corsi block Tapping Test* | 4.83 ± 1.05 |
| *ROCFT - Copy* | 32.06 ± 4.83 |
| *ROCFT - Reproduction* | 15.11 ± 6.48 |
| *RAVLT - Immediate* | 39.12 ± 9.02 |
| *RAVLT - Delayed* | 7.20 ± 3.23 |
| *Total IQ* | 90.62 ± 19.55 |
| *TMT-A* | 33.01 ± 16.30 |
| *TMT-B* | 113.48 ± 72.91 |

### 2.9. Statistics

The statistical analysis was conducted using MATLAB 2020a. To compare Iself, Iothers, and I-diff values among the three groups, a PERMANOVA test with 10,000 permutations was employed. Pairwise post-hoc comparisons were executed through permutation testing, involving the random rearrangement of labels for the two groups 10,000 times. At each iteration, the absolute value of the difference was computed, resulting in a distribution of randomly determined differences (Nichols & Holmes, 2002). This distribution was then compared to observed differences to determine statistical significance. The potential relationships between variables were explored using Pearson’s correlation and a multilinear regression model with k-fold cross-validation (Varoquaux et al., 2017). Results underwent correction through false discovery rate (FDR) correction (Benjamini & Hochberg, 1995). The significance level was established at a p-value < 0.05 after correction.

## 3. Results

We set out to investigate brain fingerprinting in epileptic patients and controls, based on the brain dynamic evaluated by the means of ATMs. Firstly, we built the differentiation matrices for each group (i.e., healthy controls, patients with left unitemporal epilepsy, patients with right unitemporal epilepsy, and patients with bitemporal epilepsy) (Figure 2A). Thereafter, we observed significant differences in fingerprinting parameters; patients with unitemporal epilepsy were grouped since they did not show significant differences among themselves, while they showed the same significant differences compared to the other groups. Then, we investigated the stability of the dynamical brain patterns in each group. Finally, we investigated possible correlations between the Iclinical score and the neuropsychological variables.

**Figure 2.**
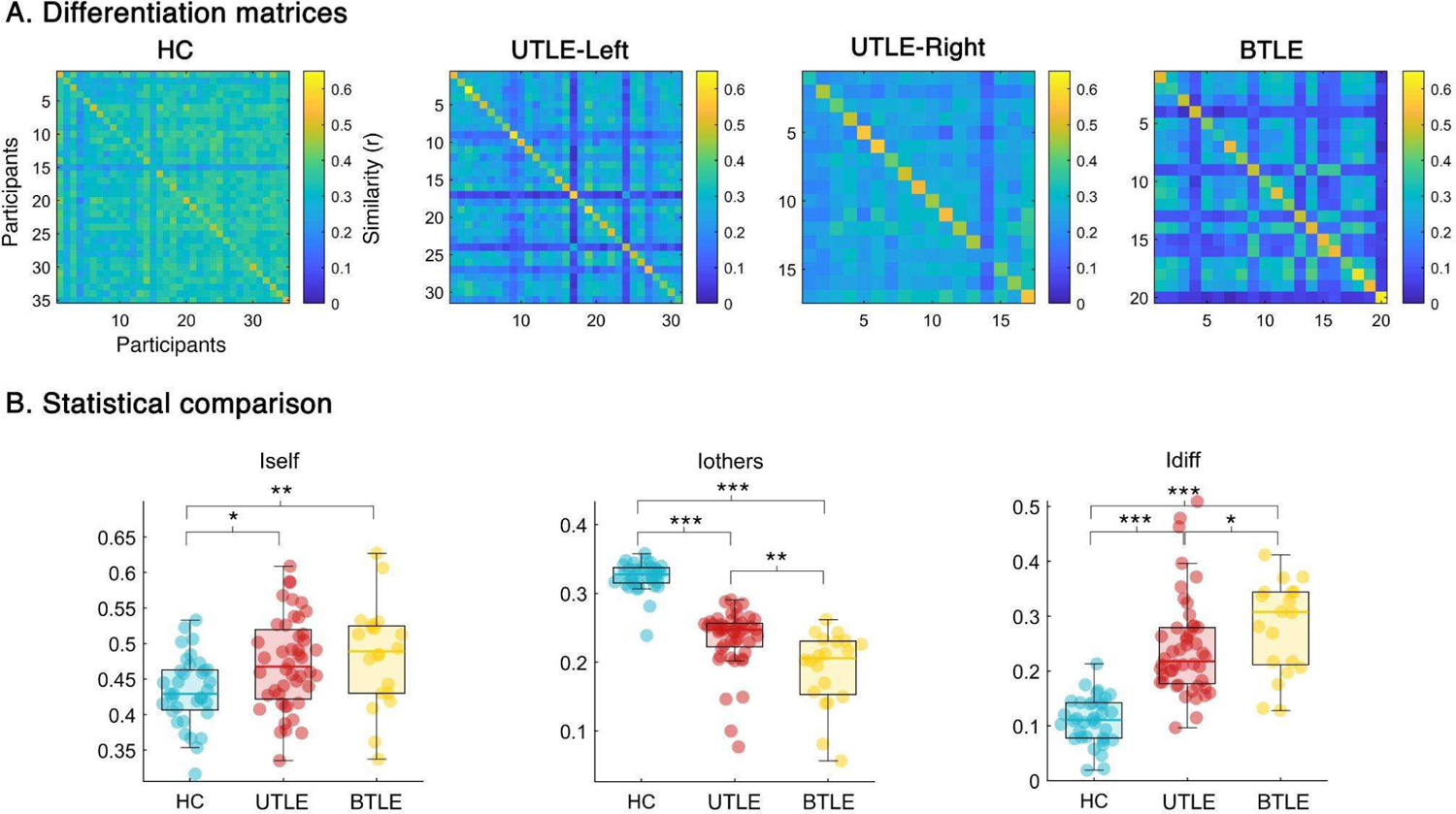
Differentiation analysis. **A** shows the differentiation matrices based on the avalanche transition matrix (ATM) of healthy controls (HC), patients with left and right unitemporal epilepsy (UTLE-Left and UTLE-Right, respectively) and patients with bitemporal epilepsy (BTLE). The matrices present participants on rows and columns, while the elements within the matrices represent the similarity (measured using Pearson correlation coefficient (r)) between test and retest ATMs of the respective individuals. **B** shows the statistical comparison among healthy controls, patients with unitemporal epilepsy (both left and right) (UTLE) and patients with bitemporal epilepsy (BTLE). Left and right UTLE patients were grouped since they did not show significant differences among themselves, while they showed the same significant differences compared to the other groups. Significance level was assessed after false discovery rate correction as follows: * < 0.05, ** < 0.01, *** < 0.001.

### Fingerprinting analysis

The PERMANOVA test (Figure 2B), performed on the Iself parameter that represents how similar two ATMs of the same individual are to each other, displayed significant differences among the three groups (p < 0.001). In particular, we found significant lower values in healthy controls with respect to patients with unitemporal (p = 0.023) and bitemporal (p = 0.006) epilepsy. Iothers resulted to be significant too (PERMANOVA test, p < 0.001). In this case, not only the healthy controls presented higher values than unitemporal (p < 0.001) and bitemporal (p < 0.001), but we also found that the unitemporal displayed higher Iothers’ values than bitemporal (p = 0.001). Finally, the analysis of the Idiff resulted as significant too (p = 0.041). We found the lowest values in healthy controls (vs UTLE, p < 0.001; vs BTLE, p < 0.001), and the higher values in BTLE (vs UTLE, p = 0.048).

### Edges’ stability

We investigated the stability of the edges of the ATMs, according to each group of participants. In line with the ICC analysis, higher ICC values corresponded to an increased stability of a given edge across the test-retest recordings of the examined group. In this case, we separated the right and left unitemporal participants, as this analysis considers the values of each specific edge across the participants. Figure 3 shows the regional contribution to the edges’ stability in the four groups. The HC displays the lowest stability globally (ICC mean in HC = 0.154, p < 0.001 vs all patients’ groups; UTLE-Left mean = 0.339, vs UTLE-Right mean = 0.267, vs BTLE mean = 0.36), suggesting that there is higher heterogeneity in the spreading patterns in physiological conditions. Conversely, patients display higher stability and this alteration is mainly distinct in the bitemporal condition. It is interesting to note the marked involvement of the temporal lobe in patients with left UTLE. In the right UTLE the corresponding lobe does not display a similar behavior, while in bitemporal condition the involvement can be mainly observed in the left lobe.

**Figure 3.**
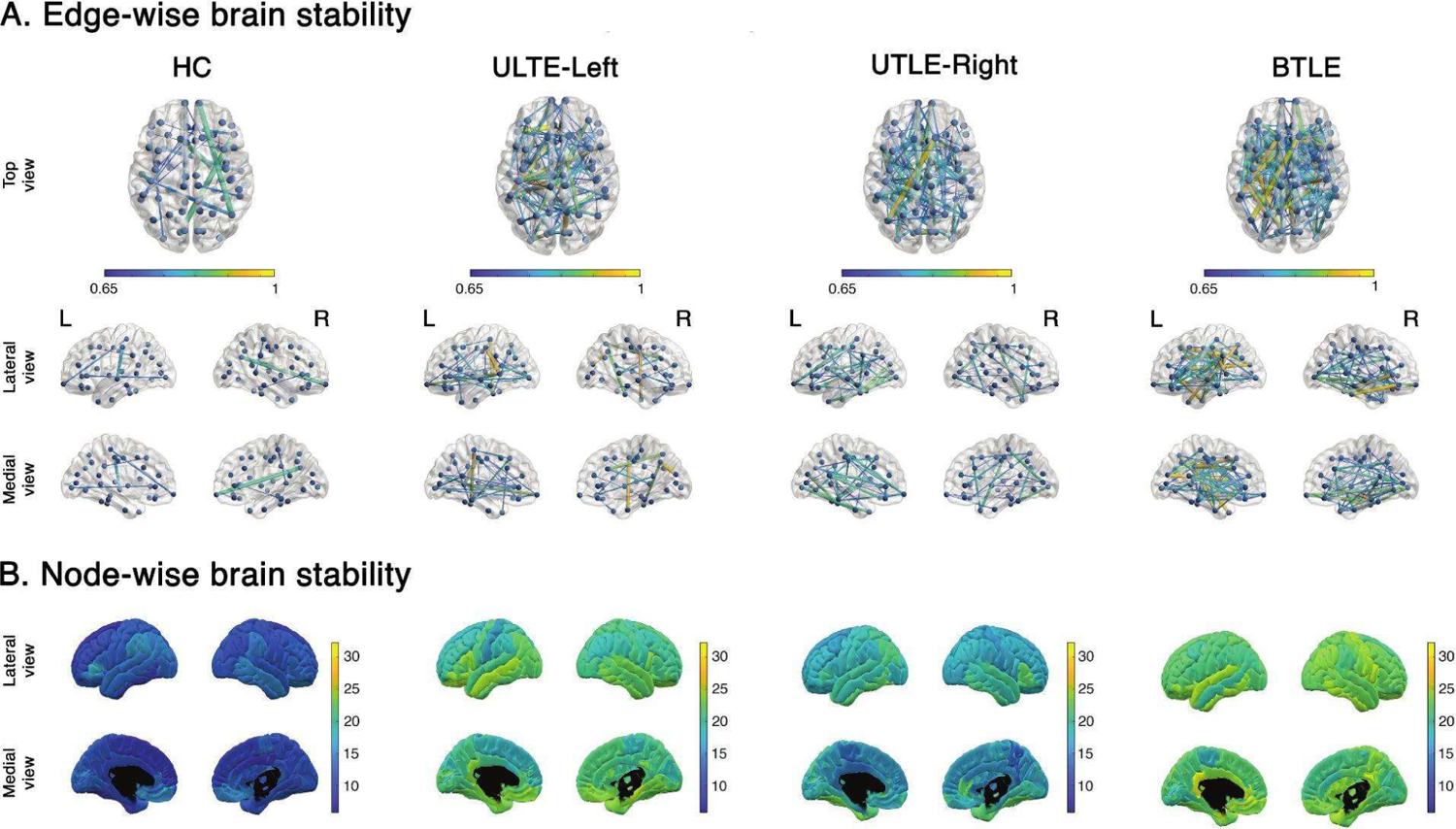
Stability of brain activity. Brain plots revealing the stability of nodes and edges in each group, obtained by the means of intraclass correlation analysis. **A.** The figure shows the edge-wise stability from the top view, and from lateral and medial views of both brain hemispheres. **B.** The figure shows the nodal stability from lateral and medial views.

### Clinical correlation

Borrowing from a previous study we calculated the Iclinical score, which represents how much the ATM of a patient resembles the average ATMs of the healthy controls. We found that, in patients with unitemporal epilepsy, the Iclinical was significantly correlated to the score of the figure recall test (ROCF-recall) (r = 0.48, p = 0.004) (Figure 4). None of the remaining clinical variables displayed significant correlations.

**Figure 4.**
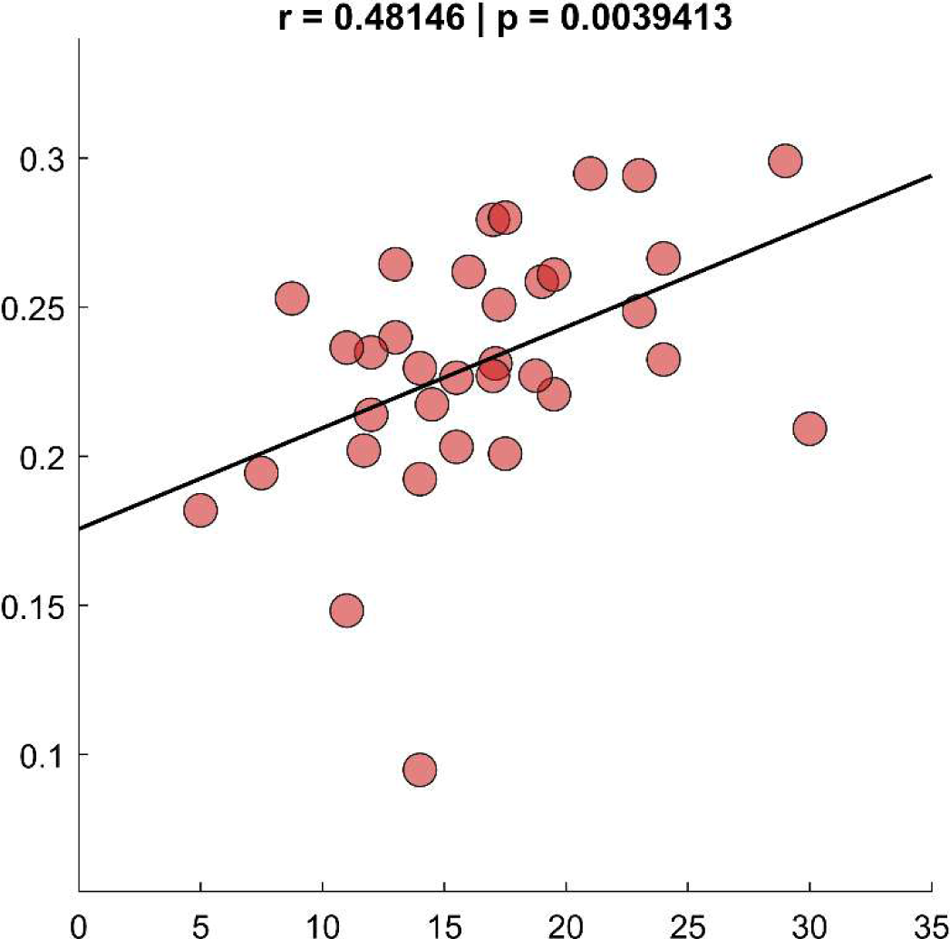
Clinical correlation. The figure shows the scatter plot of the correlation between Iclinical values and the ROCF-recall scores of the patients with unitemporal epilepsy. The more the patients were similar to the healthy controls (higher Iclinical), the better the recalling performance.

### Regression model

Furthermore, we also tested the ability of the Iclinical to predict, together with other predictors (i.e., affected hemisphere, gender, and age), the ROCF-recall scores. Hence, we built a multilinear regression model validated with a 5-fold cross validation (Figure 5) over 4000 iterations, and found that both age (β = −0.41, p =0.026) and Iclinical (β = 0.37, p = 0.041). The cross-validated model resulted to be significant (F(4,29) = 4.39, p = 0.007), with an explained variance equal to 18.8% (R2 = 0.188), a prediction error equal to 20% (NRMSE = 0.2), and a correlation coefficient between predicted and actual ROCF-recall scores equal to 0.718.

**Figure 5.**
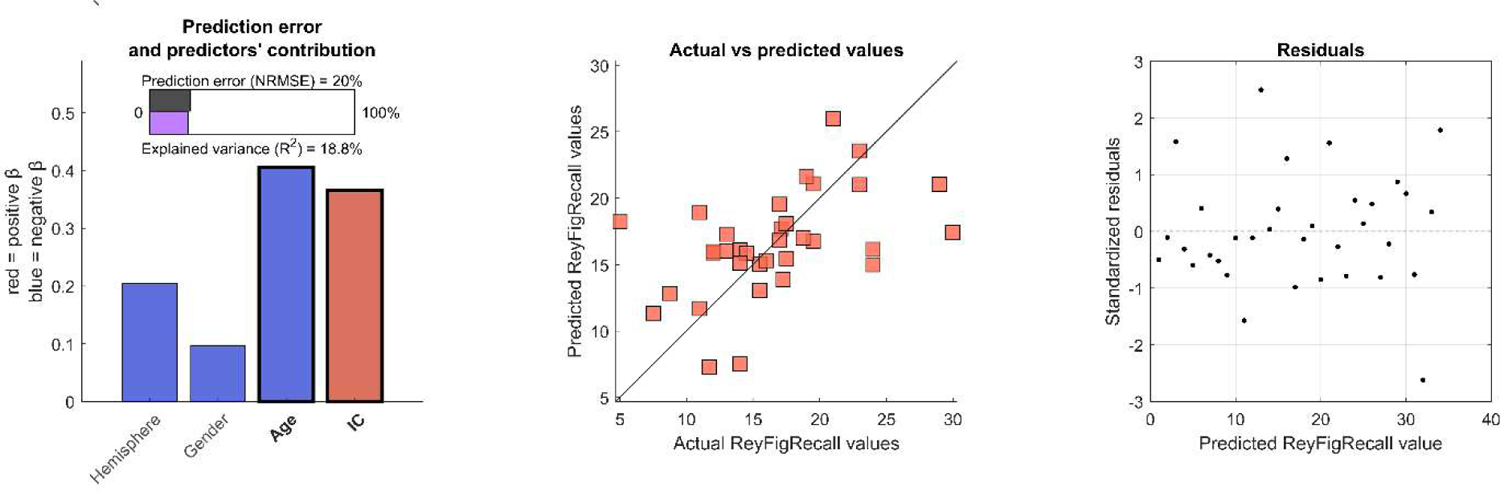
Multilinear regression model for clinical prediction. The figure shows the results of the multilinear model without (first row) and with (second row) 5-fold cross-validation. The multilinear model significantly predicts the scores of the Rey–Osterrieth complex figure recall test (ROCF-recall) in patients with unilateral epilepsy. The model is based on four predictors (i.e., lateralization of the condition, Gender, Age, Iclinical (IC). The left panels report the statistics of the model; predictors’ values are z-scored in order to make the beta coefficients comparable; significant predictors are reported in bold; NRMSE: normalized root mean square error. The middle panel is a scatter plot that compares the actual ROCF-recall scores with the ROCF-recall scores predicted by the model. The more the predictors are aligned along the diagonal, the higher is the accuracy of the prediction. Finally, the third panel shows the distribution of the standardized residuals.

## 4. Discussion

In this study, we set out to investigate whether brain dynamics may represent a neural fingerprint to identify individuals and their clinical condition, namely epilepsy. We leveraged previous findings showing that neuronal avalanches capture the altered functional organization in epilepsy (Duma et al., 2023, 2024). Recent findings highlighted that ATMs increase the performance in subject identification, i.e. neural fingerprint (Sorrentino et al., 2023). In this light, we quantified the similarity between the ATMs across and within groups, to test the hypothesis that the changes in large-scale dynamics may characterize the individual neural fingerprint and differentiate between patients and controls. While multiple studies have demonstrated altered functional configurations in TLE, we chose to focus on changes in individual patients as compared to the healthy controls to better incorporate the intra-individual variability characterizing this pathology. As a first result, we observed that patients diverge from the ‘healthy’ optimal configuration observed in controls, as they display more stereotyped dynamics. As such, each patient is more similar to him/herself over time (larger Iself values), and less similar to the other patients (reduced Iothers value). Then, we chose to analyze in-depth what edges were driving the differences in identifiability. The edge-based results provide an additional piece of information on the global dynamics of patients. The controls were characterized by more flexible brain dynamics configurations, as shown by the lower number of stable connections across brain regions, as compared to patients. Conversely, TLE patients showed more stable edges, and more stereotyped dynamics of the fronto-temporal regions. Importantly, the number of stable edges increased from unilateral to bilateral TLE (see Fig. 3). At first, these findings highlight that epilepsy is a network disorder impacting the brain dynamics at the whole-brain level. Secondly, while TLE may be considered a homogeneous clinical category, there is an array of clinical presentations according to the portion of the lobe involved in seizure generation (Bartolomei et al., 2008; Narasimhan et al., 2020; Song et al., 2022). Such heterogeneity may be mirrored in corresponding variability of brain dynamics on the large scale. Our findings, while corroborating the concept of TLE as a heterogeneous category, provide novel insight into the possibility of individual identification based on the functional reconfiguration of the brain networks, leveraging the concept of personalized medicine. Additionally, the brain dynamics organization represents a sensitive measure of the lateralization of the clinical condition. In fact, we observed decreased Iothers value in the BTLE patients, supporting a difference in the functional configuration of brain activity as compared to the UTLE. Accordingly, BTLE has been proposed as a separable and specific condition as compared to unilateral TLE (UTLE) (Chiang et al., 2022; Didato et al., 2015). Recent evidence suggests increased segregation and lower global efficiency in the functional networks of patients with BTLE (Lucas et al., 2023). Our results align with the observed altered segregation/integration ratio in BTLE, highlighting a reduction in the repertoire of brain activity reconfiguration, resulting in more stereotyped dynamics in this population. A high degree of intra-individual variability in brain activity patterns can be interpreted as an indicator of a healthy brain. This concept is based on the idea that the variability reflects the ability of the brain to flexibly adapt to multiple cognitive and behavioral tasks. To achieve this ability, the brain alternates moments of coherent activities over the large scale (integration) with moments of rearrangement of the activities (segregation) where no obvious pattern is observed on the large scale. The fine tuning of the integration-segregation ratio (Sporns, 2013) is considered to be optimizing the system capability to efficiently process environmental stimuli, while minimizing potential damage (Cohen & D’Esposito, 2016). The physiological variation in brain activity patterns reflects the complexity and uniqueness of each individual, configuring a “neural fingerprint”. However, it must be noted that quantifying the trade-off between variability/flexibility is challenging, and it can only be achieved as relative to the (presumably) optimal configuration observed in the healthy controls. In this case, brain pathology is often associated with a loss of flexibility and the emergence of stereotyped activities (Polverino et al., 2022). In neurological and psychiatric disorders, rigid and repetitive brain activity patterns are often observed, which has been related to cognitive impairment (Liang et al., 2021; Wang et al., 2022). Importantly, patients with TLE are characterized by the impairment of multiple cognitive domains, which has been linked to the dysregulation of reconfiguration properties of brain dynamics at the large scale (Caciagli et al., 2023; Girardi-Schappo et al., 2021; He et al., 2018). Our results showed that the more the patterns of propagation of the whole-brain dynamics in patients resembled those of healthy controls, the better the cognitive proficiency, in this limited to the long-term memory (recall of the ROCFT). Overall, our results corroborate the use of ATMs as a straightforward way to capture subject-specific, large-scale spatio-temporal dynamics. The ATMs have proved to be sensitive to pathology-induced alterations of brain dynamics, being able to discriminate between controls and patients with TLE. Additionally, by using ATMs we have been able to provide novel insight on the neurofunctional mechanisms distinguishing UTLE from BTLE. Moreover, our findings endow the reconfigurations of neural activity with a functional meaning since they relate to a cognitive process that has been long known to be impaired in patients with TLE, namely memory. Importantly, our results are based on a signal cleaned from epileptiform activity with twofold implications. At first from a theoretical perspective we highlighted how the basal process of regulation of neural dynamics is altered in this clinical condition. Secondly, having a pathology-sensitive metric, characterizing individual patterns of neural activity without the need of epileptiform activities, drastically increases the usability in a real clinical scenario. This opens new possibilities for a tailored investigation of brain dynamics in disease conditions, providing novel metrics for a fine-tuning of patient-specific brain models.

## Data Availability

The data that support the findings of this study are available on request to the corresponding author. The raw data are not publicly available due to privacy or ethical restrictions.

## Authors’ contributions

ETL, Conceptualization, Methodology, Software, Funding acquisition, Writing - Original Draft MCC, Conceptualization, Methodology, Formal Analysis, Writing - Original Draft AD, Investigation, Writing- Reviewing and Editing, LA, Resources, Data Collection, Writing- Reviewing and Editing MA, Software, Visualization, Writing- Reviewing and Editing PB, Investigation, Writing- Reviewing and Editing PP, Project administration, Supervision, Writing - Original Draft GMD, Project administration, Data Curation, Supervision, Writing - Original Draft, Funding acquisition

